# Investigating Unconscious Race Bias and Bias Awareness Among Vascular Surgeons

**DOI:** 10.1101/2024.06.04.24308457

**Authors:** Kerry A. Howard, Brian Witrick, Ashley Clark, Avery Morse, Karen Atkinson, Pranav Kapoor, Katharine L. McGinigle, Samantha Minc, Olamide Alabi, Caitlin W. Hicks, Andrew Gonzalez, Crystal W. Cené, Samuel Cykert, Corey A. Kalbaugh

## Abstract

**Background:** Implicit bias can influence behavior and decision-making. In clinical settings, implicit bias may influence treatment decisions and contribute to health disparities. Given documented Black-White disparities in vascular care, the purpose of this study was to examine the prevalence and degree of unconscious bias and awareness of bias among vascular surgeons treating peripheral artery disease (PAD).

**Methods:** The sampling frame included all vascular surgeons who participate in the Vascular Quality Initiative (VQI). Participants completed a survey which included demographic questions, the race implicit association test (IAT) to measure magnitude of unconscious bias, and six bias awareness questions to measure conscious bias. The magnitude of unconscious bias was no preference; or slight, moderate, or strong in the direction of pro-White or pro-Black. Data from participants were weighted to account for nonresponse bias and known differences in the characteristics of surgeons who chose to participate compared to the full registry. We stratified unconscious and conscious findings by physician race/ethnicity, physician sex, and years of experience. Finally, we examined the relationship between unconscious and conscious bias.

**Results:** There were 2,512 surgeons in the VQI registry, 304 of whom completed the survey, including getting IAT results. Most participants (71.6%) showed a pro-White bias with 73.0% of this group in the moderate and strong categories. While 77.5% of respondents showed conscious awareness of bias, of those whose conscious results showed lack of awareness, 67.8% had moderate or strong bias, compared to 55.7% for those with awareness. Bias magnitude varied based on physician race/ethnicity and years of experience. Women were more likely than men to report awareness of biases and potential impact of bias on decision-making.

**Conclusions:** Most people have some level of unconscious bias, developed from early life reinforcements, social stereotypes, and learned experiences. Regarding health disparities, however, these are important findings in a profession that takes care of patients with PAD due to heavy burden of comorbid conditions and high proportion of individuals from structurally vulnerable groups. Given the lack of association between unconscious and conscious awareness of biases, awareness may be an important first step in mitigation to minimize racial disparities in healthcare.

**CLINICAL PERSPECTIVE:** *What is new?:*

- This is the first study to examine unconscious and conscious bias in vascular surgeons, an important population for treating peripheral artery disease which disproportionately affects structurally vulnerable groups.
- We found that the majority of vascular surgeons show a pro-White bias, there is a lack of association between unconscious bias and conscious awareness of bias, and those who do not report conscious awareness of bias may also show greater magnitude of unconscious bias.

*What are the clinical implications?:*

- These findings offer important considerations for attentiveness to both unconscious biases and enhancement of awareness of the existence of biases among a surgery community that provides care to a diverse population of patients with PAD and disparities in health outcomes.
- Incorporating information on the awareness of biases and structural changes to facilitate behavior change based on these findings may be helpful within training programs for vascular surgeons.
- With awareness as an important first step in mitigation, cognizance of the existence of biases, as identified by this study, can aid in efforts to minimize racial disparities in health care.

## INTRODUCTION

Racial disparities in health care and resultant poor outcomes have been documented for patients suffering from peripheral artery disease (PAD).^1^ Patients with PAD represent a vulnerable cohort of patients with high comorbidity burden, management that requires a complex interplay of medical and interventional specialties and decision-making, and high risk for amputation and early mortality.^2–8^ Studies focused on PAD-related care delivered to Black and White patients have shown that Black patients are offered different procedures than White patients, including a lower incidence of limb-sparing revascularizations and a higher incidence of primary limb amputations (i.e., amputation without attempt at revascularization).^5^ Therefore, while all patients with PAD have some degree of risk for limb loss, Black patients with PAD have a 2-5 times greater lifetime risk of lower extremity amputation than White patients.^4^ Black patients are also less likely to receive guideline-directed medical therapy such as antiplatelet medications and lipid lowering therapy.^9^ Such disparities persist even after adjusting for variables such as disease severity, insurance status, and access to care.^10^ Taken together, such care delivery and outcome findings highlight clinical decision-making as a contributing factor to differences in care. Therefore, factors that influence decision making of vascular specialists need to be examined.

The etiology of these disparities in care is unclear, however one potential contributing factor to inequities is implicit bias, defined as an unconscious bias that may influence behavior, cognitive processes, and decision-making.^11,12^ As described in several recent systematic reviews, health care providers have implicit biases that may impact the delivery of health care.^12–15^ While there are no studies among a vascular surgeon physician population, studies have shown general physician biases in relation to care. Notably, physicians often rate Black patients more negatively (less intelligent, less compliant, etc.) than White patients.^16^ In fact, Black patients report lower ratings of care when their clinician has stronger pro-White biases, suggesting noticeable differences in the delivery of care based on bias.^17^ Furthermore, Physician awareness of biases and the potential for these to affect decision-making may be crucial towards mitigating bias and optimizing care.^11,18^ Specific to PAD, vascular surgery represents a specialty that can offer the full complement of treatments for PAD, from guideline-directed medical therapy to peripheral revascularization and lower limb amputation. Given race-based disparities present in PAD care, vascular surgeons’ implicit bias and awareness of that bias, remain an under-investigated potential factor in PAD-related Black-White treatment and outcome differences.

Given evidenced differences in care based on race, including notable differences among patients with PAD, we sought to examine the prevalence and degree of unconscious bias and awareness of bias among vascular surgeons treating PAD. We hypothesized that there would be unconscious White preferences among surgeons, as well as lack of conscious awareness of such biases.

## METHODS

### Study Sample and Source Population

The Society for Vascular Surgery (SVS) Vascular Quality Initiative (VQI) is a national vascular procedure-based registry^19^ intended to document and improve the quality of care delivered to patients with vascular diseases. All clinicians that deliver vascular care in the United States (US) are eligible to participate in the VQI. As of June 2024, the VQI includes 1,028 sites and has collected over 1.2 million procedures. The VQI has 18 regional quality groups across the US: South (5), Northeast (4), Midwest (4), and West (5). Centers include academic medical centers, teaching hospitals, community hospitals, and private practice sites.

### Recruitment strategy

We invited all 2,512 vascular surgeons who practice at VQI participating centers to take the race Implicit Association Test (IAT)^20^ in order to categorize their level of implicit bias.

Vascular surgeons who consented to participate in the study were able to take the survey on any digital device with a keyboard or a touch screen. A $100 gift card was offered as an incentive for completion of the survey. The Institutional Review Boards at Clemson University (IRB 2020- 096) and Indiana University (IRB 16325) approved this study. All data were administered and stored on a secure SSL encrypted website managed by Project Implicit, the company that created the survey.

### Survey components

The survey included three components: physician demographics, the race Implicit Association Test, and Bias Awareness questions.

#### Demographics

Survey participants were asked to provide their age, sex assigned at birth (male, female), gender identity (male, female, trans male/trans man, trans female/trans woman, genderqueer/gender nonconforming, a different identify), race, ethnicity (Hispanic, non-Hispanic, other), and years of experience (0-10, 11-20, and more than 20 years). For the purposes of this study, self-identified race and ethnicity categories were combined into Hispanic, Non-Hispanic Asian, Non-Hispanic White, Non-Hispanic Black, and Non-Hispanic Other.

#### Race Implicit Association Test

Created in 1998, the race IAT (developed by Project Implicit) is a validated instrument that measures the degree of implicit bias a participant has for individuals who are European American (White) or African American (Black).^21^ Participants are shown a series of White and Black individuals as well as ‘good’ and ‘bad’ words and are then asked to group the individuals shown with each word presented on the screen as quickly as possible. The underpinning of the IAT is that participants will match a group (images of Black or White Americans) to a characteristic, evaluation, or stereotype (‘good’ and ‘bad’ words such as excellent and angry, respectively) more quickly if there is already an implicit connection.

Differential response times (‘D scores’) are recognized as a proxy for unconscious bias. D Scores adjust for variations between innate reaction time and calculate the difference in reaction time between two blocks of the trial. A positive score indicates pro-White bias while a negative score indicates pro-Black bias. Scores include: -0.15 to 0.15 (no bias), 0.16 to 0.34 (slight bias), 0.35 to 0.64 (moderate bias), and > 0.65 (strong bias).

#### Bias Awareness Measure

Following completion of the IAT, six self-administered survey questions were used to measure participant conscious bias: “I am biased,” “Everyone, including me, has biases toward other people,” “Unwanted biases probably influence my decisions about other people,” “Unwanted bias may affect the way I make decisions without my realizing it,” “Unwanted biases may influence my perception of the world around me,” and “Unwanted bias may influence my interactions with other people.” Each question included strongly agree, moderately agree, slightly agree, slightly disagree, moderately disagree, strongly disagree, and prefer not to answer as response options.

### Statistical analysis

In order to reduce the potential for nonresponse bias and effect of known differences in the characteristics of surgeons from the VQI registry who chose to participate in this study compared to the full registry, data were weighted prior to analysis. First, we cross-classified physicians into “weighting cells” based on the following three variables: race/ethnicity, age category, and sex. These variables were chosen because there were differences in the likelihood of participating in the study and in study outcomes based on these variables.^22^ Then, a weight defined as the number of physicians on the registry divided by the number of physicians who participated in the study was applied within each weighting cell (for example, male physicians 55+ years of age who identified as non-Hispanic White made up one weighting cell). This adjustment effectively “weights up” physicians from demographic groups who were underrepresented in the data and “weights down” physicians from groups that were overrepresented.

Based on IAT results, physicians were grouped according to one of seven categories based on degree of implicit bias: no bias, slight bias toward White, moderate bias toward White, strong bias toward White, slight bias toward Black, moderate bias toward Black, and strong bias toward Black. We also examined both IAT results and conscious bias awareness results by strata of physician race/ethnicity, physician sex, and physician years of experience. In these analyses, IAT results were grouped in two ways: 1) pro-White bias, pro-Black bias, and no bias; and 2) four categories based on the magnitude of the association: no bias, slight bias, moderate bias, and strong bias. Bias awareness results were grouped into three categories based on the majority of a participant’s answers: generally agree, generally disagree, and a combination of agree and disagree. Chi-square analyses tested the distribution of bias results across strata. We used Spearman Correlation to test for an association between implicit bias magnitude and bias awareness, as well as chi-square for the distribution of implicit bias results across two levels of awareness: average of aware answers and average of unaware answers. All analyses were run using SAS software version 9.4 (SAS Institute, Cary, NC) survey procedures that reflect weighting in the computation of estimates.

## RESULTS

Among 2,512 VQI-participating vascular surgeons, 368 (14.6%) physicians started the survey and 304 (12.1%) physicians completed the entire survey, including obtaining results from the IAT. The average age of participants was 44 years. The majority of the sample were Non-Hispanic White (61.8%) and male (66.8%). While the survey asked about both sex and gender and provided options other than sex assigned at birth for gender, all respondents selected their sex assigned at birth as their gender. The ethnoracial distribution of participants included Non-Hispanic Black (4.6%), Hispanic (4.6%), Non-Hispanic Asian (23.0%), and individuals who identified as another non-Hispanic race (5.9%). As described above, weights were applied to account for the differences in our sample and the VQI frame. Sample characteristics and the weighted values based on the full VQI frame are provided in Table 1.

**Table 1.** Samples characteristics and weighted values based on characteristics of the Vascular Quality Initiative frame.

| Physician Characteristics | Sample | Weighted Value |
| --- | --- | --- |
| Age, Mean (standard deviation) | 44.3 (10.1) | 50.6 (10.8) |
| Race/Ethnicity, n (%) |  |  |
| Hispanic | 14 (4.6) | 14 (4.5) |
| Non-Hispanic Asian | 70 (23.0) | 52 (17.2) |
| Non-Hispanic Black | 14 (4.6) | 11 (3.7) |
| Non-Hispanic Other | 18 (5.9) | 18 (6.1) |
| Non-Hispanic White | 188 (61.8) | 209 (68.7) |
| Sex, n (%) |  |  |
| Male | 203 (66.8) | 272 (89.5) |
| Female | 101 (33.2) | 32 (10.5) |
| Years of Experience, n (%) |  |  |
| 0-10 years | 189 (62.2) | 103 (33.8) |
| 11-20 years | 57 (18.8) | 84 (27.7) |
| More than 20 years | 58 (19.1) | 117 (38.5) |

Frequencies (n) for weighted values are rounded to the nearest whole number to indicate the number of persons

### Overall Implicit Bias and Awareness Results

We found that 18.8% (n=57) of vascular surgeons had no bias for Black or White individuals. The majority, 71.6% (n=218), of physicians had a pro-White bias and 9.7% (n=29) had a pro-Black bias. Of the approximately (due to weighting) 218 vascular surgeons with a pro-White bias, 27.0% were found to have a slight bias, 45.3% had a moderate bias, and 27.7% had a strong bias for White compared to Black individuals. Of the approximately 29 surgeons with a pro-Black bias, 36.3% were found to have a slight bias, 52.7% had a moderate bias, and 11.0% had a strong bias for Black compared to White individuals. Overall, 39.2% tended to show strong awareness of biases and the potential for biases to affect decision-making, while 10.3% tended to strongly indicate lack of awareness, and 50.5% gave mixed responses. Based on the average of a person’s responses, 77.5% generally showed awareness and 22.5% generally showed lack of awareness. Pro-White bias, pro-Black bias, and awareness results stratified by physician race/ethnicity, physician sex, and physician years of experience are presented below and in Table 2.

**Table 2.** Distribution of implicit bias and awareness of bias results by physician characteristics.

| Physician Characteristics | No preference | Pro-White Bias |  |  | Pro-Black Bias |  |  | Awareness |  |
| --- | --- | --- | --- | --- | --- | --- | --- | --- | --- |
|  |  | Slight | Moderate | Strong | Slight | Moderate | Strong | Unaware | Aware |
| Race/Ethnicity, n (%) |  |  |  |  |  |  |  |  |  |
| Hispanic | 2 (16.0) | 1 (4.6) | 8 (59.9) | – | 1 (4.63) | 2 (14.8) | – | 4 (28.7) | 10 (71.3) |
| Non-Hispanic Asian | 12 (22.2) | 11 (20.3) | 19 (36.7) | 9 (18.0) | 1 (2.2) | 0 (0.6) | – | 10 (19.1) | 42 (80.9) |
| Non-Hispanic Black | 5 (40.8) | 2 (19.6) | 1 (12.8) | – | 0 (1.5) | 2 (18.1) | 1 (7.2) | 1 (11.3) | 10 (88.7) |
| Non-Hispanic Other | 2 (11.8) | 9 (50.8) | 2 (11.2) | 2 (11.8) | – | 3 (14.3) | – | 4 (21.2) | 14 (78.8) |
| Non-Hispanic White | 37 (17.5) | 36 (17.2) | 68 (32.5) | 49 (23.3) | 9 (4.2) | 8 (4.1) | 2 (1.2) | 49 (23.7) | 159 (76.3) |
| Sex, n (%) |  |  |  |  |  |  |  |  |  |
| Male | 51 (18.9) | 53 (19.5) | 86 (31.8) | 56 (20.5) | 8 (2.9) | 15 (5.4) | 3 (1.1) | 65 (23.9) | 207 (76.1) |
| Female | 6 (17.6) | 6 (18.1) | 12 (37.8) | 4 (14.1) | 3 (9.2) | 1 (2.6) | 0 (0.5) | 4 (10.9) | 28 (89.1) |
| Years of Experience, n (%) |  |  |  |  |  |  |  |  |  |
| 0-10 years | 22 (21.5) | 16 (15.4) | 31 (30.0) | 24 (22.9) | 3 (3.3) | 6 (6.1) | 1 (0.8) | 30 (29.2) | 73 (70.8) |
| 11-20 years | 20 (23.3) | 24 (28.9) | 30 (35.5) | 5 (5.7) | 2 (2.9) | 3 (3.8) | – | 20 (23.3) | 64 (76.7) |
| More than 20 years | 15 (13.1) | 19 (15.9) | 38 (32.3) | 32 (27.3) | 5 (4.2) | 6 (5.2) | 2 (2.1) | 19 (16.1) | 98 (83.9) |

Frequencies (n) are rounded to the nearest whole number to indicate the number of persons

### Bias and Awareness by Physician Race and Ethnicity

Based on results of the IAT, the majority of Hispanic (64.5%), Non-Hispanic Asian (75.0%), and Non-Hispanic White (73.1%) physicians had an unconscious pro-White bias. Non-Hispanic Black physicians had a more even distribution across pro-White (32.5%), pro-Black (26.8%), and no bias (40.8%). There was a statistically significant difference in the distribution of IAT result magnitude (none, slight, moderate, strong) across physician race/ethnicity categories (ꭓ2 = 47.44, p =.003). Black physicians were most likely to have no bias (40.8%), followed by a moderate bias (31.0%), slight bias (21.1%) and strong bias (7.8%). Hispanic (74.7%), Non-Hispanic Asian (37.2%), and White (36.6%) physicians were most likely to have moderate bias. The majority of physicians identifying as Non-Hispanic Other had a slight bias (50.8%). Strong bias was the least likely finding for all races/ethnicities except White, for whom it was second most likely (24.5%). Non-Hispanic White physicians were also least likely of physician race/ethnic categories to have no bias (17.5%). There was no evidence of a difference in the distribution of conscious bias awareness results by physician race/ethnicity (ꭓ2 = 8.03, p =.431).

### Bias and Awareness by Physician Sex

Male (71.7%) and female (70.1%) vascular surgeons were both highly likely to have a pro-White bias. We found no detectable differences in conscious bias magnitude by surgeon sex (ꭓ^2^ = 1.10, *p* =.777). Male and female surgeons were similarly likely to have no bias (18.9% vs. 17.6%), slight bias (22.3% vs. 27.4%), moderate bias (37.2% vs. 40.4%), and strong bias (21.6% vs. 14.6%). There was a difference in awareness of bias (ꭓ^2^ = 6.22, *p* =.044) by surgeon sex.

Female vascular surgeons (89.1%) were more likely than male vascular surgeons (76.1%) to generally agree with the bias awareness questions, indicating greater awareness of the existence of bias and the effect of bias on decision-making.

### Bias and Awareness by Physician Years of Experience

The results showed a statistically significant difference in magnitude of implicit bias by physician years of experience (ꭓ^2^ = 24.36, *p* =.018). Physicians, regardless of years of experience, were most likely to have a moderate bias (0-10 years: 36.1%; 11-20 years: 39.3%; more than 20 years: 37.5%). Those with more than 20 years of experience showed less likelihood of no bias (13.1%) compared to 0-10 years (21.5%) and 11-20 years (23.3%). Those with 11-20 years of experience showed least likelihood of strong bias (5.7%) compared to 0-10 (23.7%) and more than 20 years (29.4%). There was no evidence of a difference in conscious bias awareness between groups of years of experience (ꭓ^2^ = 4.82, *p* =.307).

### Association Between Implicit Bias and Bias Awareness

We examined the association between magnitude of implicit bias (none, slight, moderate, strong) and awareness of bias through conscious bias responses. Overall, there was no evidence of an association (*ρ* = -0.03, *p* = .600). There was a trend in which those with lack of awareness of bias had a higher percent of moderate and strong implicit bias results than those with awareness (ꭓ^2^ = 3.19, *p* =.074). Of those whose conscious bias results showed lack of awareness, 67.8% had moderate or strong bias, compared to 55.7% for those with awareness. Among levels of race/ethnicity (Figure 1A), physician sex (Figure 1B), and years of experience (Figure 1C), this trend was particularly the case for physicians with more than 20 years of experience (aware, moderate or strong: 62.1%; unaware, moderate or strong: 91.5%; ꭓ2 = 6.12, p =.013) and for male physicians (aware, moderate or strong: 55.6%; unaware, moderate or strong: 69.0%; ꭓ2 = 3.69, p =.055).

**Figure 1.**
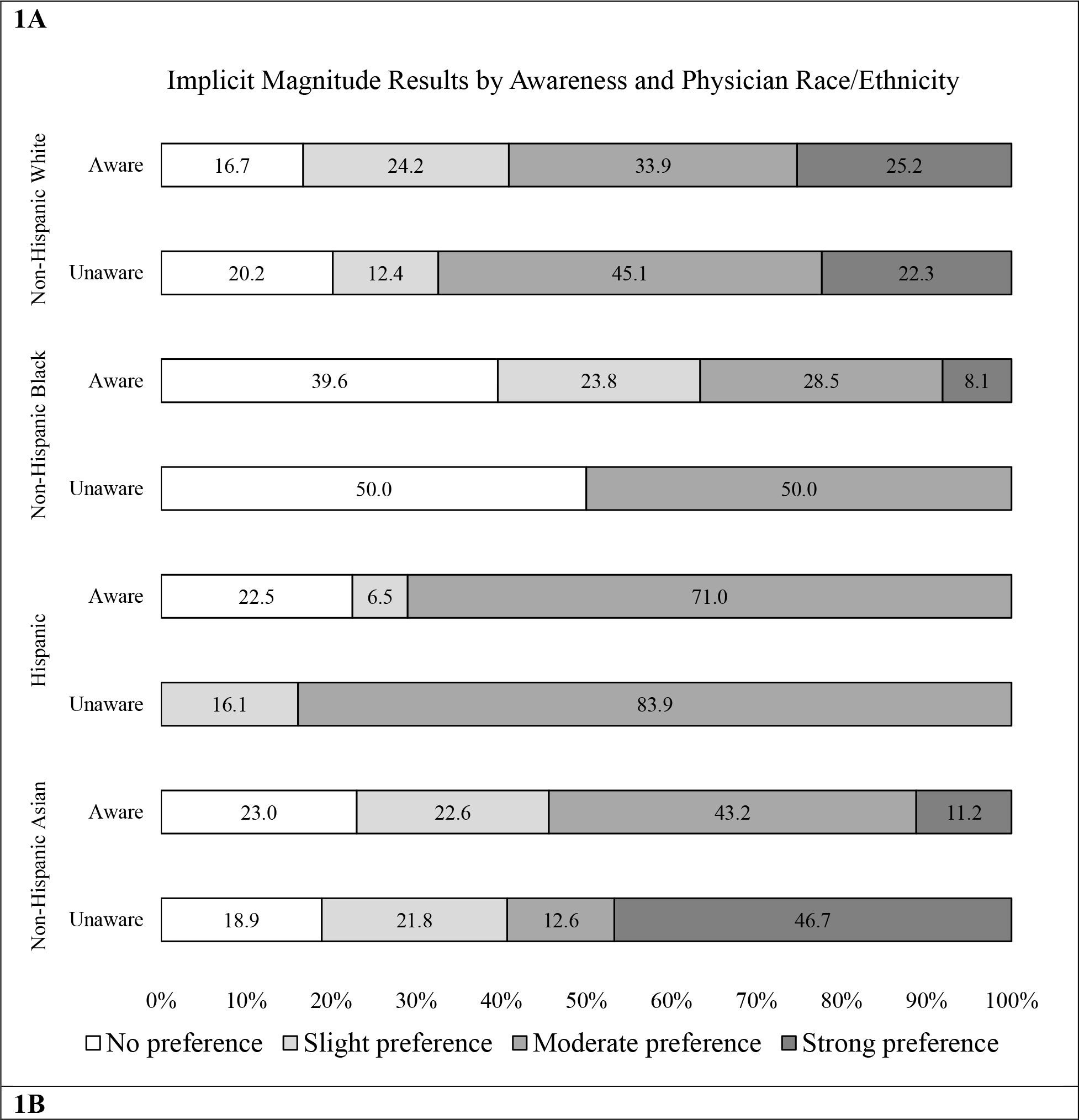
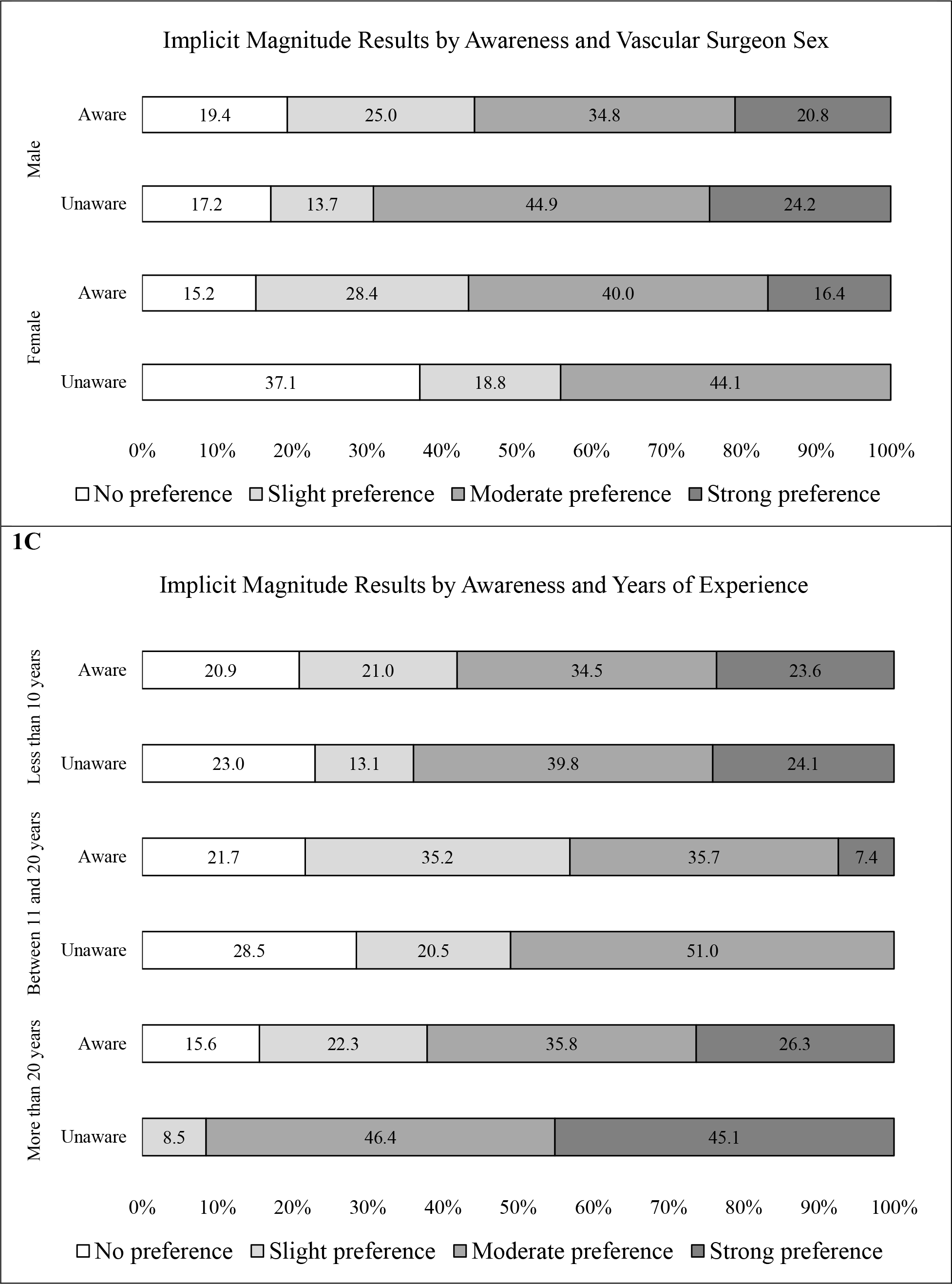
Magnitude of unconscious results by conscious awareness results. The results of the implicit association test (no bias, slight bias, moderate bias, strong bias) between unaware and aware conscious bias answers based on levels of physician race/ethnicity (1A), physician sex (1B), and physician years of experience (1C).

## DISCUSSION

The purpose of this study was to explore and describe the level of unconscious bias and conscious bias among vascular surgeons that contribute to a national quality registry. Consistent with our hypothesis, we found that the majority of vascular surgeons had some degree of pro-White bias with levels of bias that are similar to those found in the general population and in numerous health care fields.^18,23^ Important differences in unconscious bias were identified by physician race/ethnicity and physician years of experience. Non-Hispanic White, Hispanic, and Non-Hispanic Asian physicians had similarly high levels of pro-White bias. There was no evidence of a relationship between magnitude of unconscious bias and conscious bias responses. However, we found that female vascular surgeons were more likely than male vascular surgeons to generally agree that biases exist and may impact decision-making. These findings offer important considerations for attentiveness to both unconscious biases and enhancement of awareness of the existence of biases among a surgery community that provides care to a diverse population of patients with PAD and disparities in health outcomes.

The majority of people have some level of unconscious bias: an analysis of 14 years of IAT data found that 65% of respondents showed a pro-White bias.^24^ Unconscious bias is developed from early life reinforcements, social stereotypes, and learned experiences from oneself and those around us.^14,23,25^ Bias is not always negative, as unconscious pattern recognition is key for making rapid and efficient decisions that can be necessary for survival (e.g., identifying danger) and healthcare (e.g., treating acutely ill patients).^11^ Within the intersection of healthcare and race, however, significant problems may results from implicit bias associated with negative stereotyping of individuals who identify as a minoritized race or ethnicity which, in turn, can impact decision-making. Our finding that 71.6% of vascular surgeons have a pro-White bias, while relatively consistent with the general population, is particularly noteworthy given the high prevalence of vascular diseases in Black patients as well as the poorer outcomes in Black patients with PAD compared to White patients.

Previous research has shown that conscious biases via awareness and unconscious biases are often dissociated from each other.^26,27^ For example, physicians have been found to view Black patients as non-compliant and less cooperative with care, without explicitly reporting such perceptions.^27^ Our results were consistent with these results as we did not identify a relationship between unconscious bias magnitude and conscious bias awareness responses. In fact, 67.8% of those who indicated that they are not aware of, or admit to, the existence of biases and their role in decision-making showed moderate or strong unconscious bias, compared to 55.68% among those who indicated awareness. While more overt discrimination is easier to identify, unconscious bias is present whether it is realized or not. Therefore, an arguably more pressing issue than the existence of unconscious biases is lack of awareness of one’s own bias and the potential need to address or manage bias.^11,28^ When examining trends in moderate and strong biases in our study, specifically, there was some evidence that those who report lack of awareness are more likely to have moderate or strong bias. Therefore, awareness is a critical first step in mitigation efforts.^29^ Bias training programs have been developed with mixed effectiveness for mitigating the impact of unconscious biases in clinical settings.^11^ The findings of this study provide preliminary information of the need for interventions to address awareness.^30^

Specifically, the present study offers practical implications based on stratification across physician race/ethnicity, sex, and years of experience. First, our analysis showed that those with 0-10 years of experience and those with 20 or more years of experience had a similar frequency of strong magnitude of bias. Given that vascular surgery training follows a group apprenticeship model, vascular surgery trainees may be subject to a variety of behaviors that may shape their own bias.^31^ Incorporating information on the awareness of biases and structural changes to facilitate behavior change based on this information may be helpful within training programs.^32,33^ Second, differences in unconscious biases based on physician race/ethnicity and in conscious biases based on physician sex offer opportunities for programs to address awareness on a personal level, targeting training and evaluation based on individualized strategies. Further, similar to our results, studies that seek to understand differences in implicit bias by the sex of the physician have found no observable difference in bias by sex.^13,14,34^ That said, in also examining conscious bias, we did find that female vascular surgeons reported more awareness of the existence of biases than male vascular surgeons. Diversification in the physician workforce, such that there are more practicing female vascular surgeons, may be a part of elevation of an awareness of biases which, along with other strategies discussed here, may aid in the collective goal to minimize racial health disparities.

People with PAD are a vulnerable cohort with high risk of poor outcomes, such as amputation and early mortality. While all people with PAD are at a high risk of poor outcomes, patients from minoritized communities, such as Black Americans, represent a structurally vulnerable group whose PAD-related care may be a result of intersecting political, social, and cultural hierarchies, as opposed to individual-level behaviors.^35^ Such hierarchies may have created a clinical environment where surgeons, in this case, as well as the general population, are influenced by pro-White bias. Given that factors that could reasonably explain disparities, such as disease severity and access to care, are examined and disparities remain, it is critical to consider the ways that structural issues may influence decision-making. Furthermore, given that an individual’s implicit biases are outside of their conscious awareness,^12^ issues of bias may be further compounded by a physicians’ lack of awareness. While studies have examined whether or not physicians admit to explicit biases,^36,37^ intervention for implicit bias requires understanding the extent of awareness of one’s own bias, particularly among physicians tasked with managing care for structurally vulnerable patients with PAD.

Our study has limitations and strengths that are important to discuss. The IAT was developed in the field of psychology where there is an ongoing debate over its appropriateness in measuring implicit bias.^38^ There are also questions about the numeric cut points used to delineate the magnitude of bias including if they are useful in determining the severity of the test-takers bias.^39^ However, despite these concerns, the IAT remains the most widely used test for studying unconscious bias with good reliability and validity and is superior to self-reported measures of bias.^21,40^ Additionally, our study population only included vascular surgeons that participate in the VQI, which are physicians who are part of larger hospital centers that have a focus on quality improvement in patient care. These surgeons may not be representative of all US-based vascular surgeons. Further, the sample of individuals who completed the study survey included more women and younger surgeons than the overall frame. Because we anticipated the aforementioned challenges, we were able to compare our sample to the overall frame and provide weighting to account for non-response and demographic differences in the final participating sample. Thus, we still have the ability to make inferences to the broader VQI surgeon population. That said, there may be other variables that we were unable to correct for in our weighting that could bias estimates, and a larger sample size may have been able to uncover statistically significant trends that we are unable to discern with the present sample. Additionally, self-reported measures of bias awareness may be subject to social desirability effects, as is true for many survey measures where participants may not want to report information that will paint them in a negative light. Nonetheless, our study represents an important step forward in understanding potential mechanisms that underlie racial disparities in vascular care.

## CONCLUSION

The present study provides evidence that pro-White bias is present in a significant proportion of vascular surgeons in our sample. While these findings are similar to what has been observed in studies of the general population and of medical providers in other specialties, these are important findings in a profession that takes care of patients with PAD due to a heavy burden of chronic comorbid conditions and a high proportion of individuals from structurally vulnerable groups. Given a lack of relationship between such unconscious biases and conscious awareness of biases, and their potential to impact on clinical outcomes, these findings offer considerations for delivering care to structurally vulnerable patients, such as those with PAD. With awareness as an important first step in mitigation, cognizance of the existence of biases, as identified by this study, can aid in efforts to minimize racial disparities in health care.

## Data Availability

Because of the sensitive nature of the data collected for this study, requests to access the dataset from qualified researchers trained in human subject confidentiality protocols may be sent to the Vascular Quality Initiative.

## Acknowledgements

The authors thank the staff and participants of the VQI for their important contributions. We also thank the Society for Vascular Surgery Patient Safety Organization for their support of this study from conception through completion.

## Funding Sources

CAK was funded for this work by Career Development Awards from the American Heart Association (19CDA34760135) and National Institute of Health/National Heart Lung and Blood Institute (K01HL146900).

## Disclosures

None

## Notes

### Competing Interest Statement

The authors have declared no competing interest.

### Author Declarations

The Institutional Review Boards at Clemson University (IRB 2020-096) and Indiana University (IRB 16325) approved this study.

## REFERENCES

1. Institute of Medicine (US). Unequal Treatment: Confronting Racial and Ethnic Disparities in Health Care. (Smedley B, Stith A, Nelson A, eds.). National Academies Press; 2003:12875. doi:10.17226/12875

2. Vart P, Coresh J, Kwak L, Ballew SH, Heiss G, Matsushita K. Socioeconomic Status and Incidence of Hospitalization With Lower-Extremity Peripheral Artery Disease: Atherosclerosis Risk in Communities Study. J Am Heart Assoc. 2017;6(8):e004995. doi:10.1161/JAHA.116.004995

3. Arya S, Binney Z, Khakharia A, et al. Race and Socioeconomic Status Independently Affect Risk of Major Amputation in Peripheral Artery Disease. J Am Heart Assoc. 2018;7(2):e007425. doi:10.1161/JAHA.117.007425

4. Newhall K, Spangler E, Dzebisashvili N, Goodman DC, Goodney P. Amputation Rates for Patients with Diabetes and Peripheral Arterial Disease: The Effects of Race and Region. Ann Vasc Surg. 2016;30:292–298.e1. doi:10.1016/j.avsg.2015.07.040

5. Holman KH, Henke PK, Dimick JB, Birkmeyer JD. Racial disparities in the use of revascularization before leg amputation in Medicare patients. J Vasc Surg. 2011;54(2):420–426.e1. doi:10.1016/j.jvs.2011.02.035

6. Jones WS, Patel MR, Dai D, et al. High mortality risks after major lower extremity amputation in Medicare patients with peripheral artery disease. Am Heart J. 2013;165(5):809–815.e1. doi:10.1016/j.ahj.2012.12.002

7. Swaminathan A, Vemulapalli S, Patel M, Jones S. Lower extremity amputation in peripheral artery disease: improving patient outcomes. Vasc Health Risk Manag. Published online July 2014:417. doi:10.2147/VHRM.S50588

8. Witrick B, Shi L, Mayo R, Hendricks B, Kalbaugh CA. The association between socioeconomic distress communities index and amputation among patients with peripheral artery disease. Front Cardiovasc Med. 2022;9:1021692. doi:10.3389/fcvm.2022.1021692

9. Kalbaugh CA, Witrick B, Howard KA, et al. Investigating the impact of suboptimal prescription of preoperative antiplatelets and statins on race and ethnicity-related disparities in major limb amputation. Vasc Med. 2024;29(1):17–25. doi:10.1177/1358863X231196139

10. Kalbaugh CA, Witrick B, Sivaraj LB, et al. Non-Hispanic Black and Hispanic Patients Have Worse Outcomes Than White Patients Within Similar Stages of Peripheral Artery Disease. J Am Heart Assoc. 2022;11(1):e023396. doi:10.1161/JAHA.121.023396

11. Gopal DP, Chetty U, O’Donnell P, Gajria C, Blackadder-Weinstein J. Implicit bias in healthcare: clinical practice, research and decision making. Future Healthc J. 2021;8(1):40–48. doi:10.7861/fhj.2020-0233

12. FitzGerald C, Hurst S. Implicit bias in healthcare professionals: a systematic review. BMC Med Ethics. 2017;18(1):19. doi:10.1186/s12910-017-0179-8

13. Haider AH, Schneider EB, Sriram N, et al. Unconscious Race and Social Class Bias Among Acute Care Surgical Clinicians and Clinical Treatment Decisions. JAMA Surg. 2015;150(5):457. doi:10.1001/jamasurg.2014.4038

14. Maina IW, Belton TD, Ginzberg S, Singh A, Johnson TJ. A decade of studying implicit racial/ethnic bias in healthcare providers using the implicit association test. Soc Sci Med. 2018;199:219–229. doi:10.1016/j.socscimed.2017.05.009

15. Dehon E, Weiss N, Jones J, Faulconer W, Hinton E, Sterling S. A Systematic Review of the Impact of Physician Implicit Racial Bias on Clinical Decision Making. Choo EK, ed. Acad Emerg Med. 2017;24(8):895–904. doi:10.1111/acem.13214

16. Van Ryn M, Burke J. The effect of patient race and socio-economic status on physicians’ perceptions of patients. Soc Sci Med. 2000;50(6):813–828. doi:10.1016/S0277-9536(99)00338-X

17. Blair IV, Steiner JF, Fairclough DL, et al. Clinicians’ Implicit Ethnic/Racial Bias and Perceptions of Care Among Black and Latino Patients. Ann Fam Med. 2013;11(1):43–52. doi:10.1370/afm.1442

18. Chapman EN, Kaatz A, Carnes M. Physicians and Implicit Bias: How Doctors May Unwittingly Perpetuate Health Care Disparities. J Gen Intern Med. 2013;28(11):1504–1510. doi:10.1007/s11606-013-2441-1

19. Cronenwett JL, Kraiss LW, Cambria RP. The Society for Vascular Surgery Vascular Quality Initiative. J Vasc Surg. 2012;55(5):1529–1537. doi:10.1016/j.jvs.2012.03.016

20. Greenwald AG, McGhee DE, Schwartz JLK. Measuring individual differences in implicit cognition: The implicit association test. J Pers Soc Psychol. 1998;74(6):1464–1480. doi:10.1037/0022-3514.74.6.1464

21. Greenwald AG, Poehlman TA, Uhlmann EL, Banaji MR. Understanding and using the Implicit Association Test: III. Meta-analysis of predictive validity. J Pers Soc Psychol. 2009;97(1):17–41. doi:10.1037/a0015575

22. Little RJ, Vartivarian S. Does weighting for nonresponse increase the variance of survey means? Surv Methodol. 2005;31(2):161–168.

23. Nosek BA, Smyth FL, Hansen JJ, et al. Pervasiveness and correlates of implicit attitudes and stereotypes. Eur Rev Soc Psychol. 2007;18(1):36–88. doi:10.1080/10463280701489053

24. Morehouse KN, Banaji MR. The Science of Implicit Race Bias: Evidence from the Implicit Association Test. Daedalus. 2024;153(1):21–50. doi:10.1162/daed_a_02047

25. Baron AS, Banaji MR. The Development of Implicit Attitudes: Evidence of Race Evaluations From Ages 6 and 10 and Adulthood. Psychol Sci. 2006;17(1):53–58. doi:10.1111/j.1467-9280.2005.01664.x

26. Nosek BA, Banaji MR, Greenwald AG. Harvesting implicit group attitudes and beliefs from a demonstration web site. Group Dyn Theory Res Pract. 2002;6(1):101–115. doi:10.1037/1089-2699.6.1.101

27. Green AR, Carney DR, Pallin DJ, et al. Implicit Bias among Physicians and its Prediction of Thrombolysis Decisions for Black and White Patients. J Gen Intern Med. 2007;22(9):1231–1238. doi:10.1007/s11606-007-0258-5

28. Sherman MD, Ricco J, Nelson SC, Nezhad SJ, Prasad S. Implicit Bias Training in Residency Program: Aiming for Enduring Effects. Fam Med. 2019;51(8):677–681. doi:10.22454/FamMed.2019.947255

29. Zeidan AJ, Khatri UG, Aysola J, et al. Implicit Bias Education and Emergency Medicine Training: Step One? Awareness. Runde DP, ed. AEM Educ Train. 2019;3(1):81–85. doi:10.1002/aet2.10124

30. Hicks CW. Atherectomy overuse is a real problem. J Vasc Surg. 2022;76(3):786–787. doi:10.1016/j.jvs.2022.04.026

31. Hafferty FW. Beyond curriculum reform: confronting medicine’s hidden curriculum. Acad Med. 1998;73(4):403–407. doi:10.1097/00001888-199804000-00013

32. Carter ER, Onyeador IN, Lewis NA. Developing & delivering effective anti-bias training: Challenges & recommendations. Behav Sci Policy. 2020;6(1):57–70. doi:10.1177/237946152000600106

33. DAnnibale K, Russ AC, Tierney RT, Mansell JL. Anti-racism training for healthcare professionals: A critically appraised topic. CommonHealth. 2022;3(1):28–33. doi:10.15367/ch.v3i1.498

34. Sabin JA, Nosek BA, Greenwald AG, Rivara FP. Physicians’ Implicit and Explicit Attitudes About Race by MD Race, Ethnicity, and Gender. J Health Care Poor Underserved. 2009;20(3):896–913. doi:10.1353/hpu.0.0185

35. Bourgois P, Holmes SM, Sue K, Quesada J. Structural Vulnerability: Operationalizing the Concept to Address Health Disparities in Clinical Care. Acad Med. 2017;92(3):299–307. doi:10.1097/ACM.0000000000001294

36. Qian MK, Heyman GD, Quinn PC, Fu G, Lee K. Differential developmental courses of implicit and explicit biases for different other-race classes. Dev Psychol. 2019;55(7):1440–1452. doi:10.1037/dev0000730

37. Feldner HA, VanPuymbrouck L, Friedman C. Explicit and implicit disability attitudes of occupational and physical therapy assistants. Disabil Health J. 2022;15(1):101217. doi:10.1016/j.dhjo.2021.101217

38. Oswald FL, Mitchell G, Blanton H, Jaccard J, Tetlock PE. Predicting ethnic and racial discrimination: A meta-analysis of IAT criterion studies. J Pers Soc Psychol. 2013;105(2):171–192. doi:10.1037/a0032734

39. Blanton H, Jaccard J. Arbitrary metrics in psychology. Am Psychol. 2006;61(1):27–41. doi:10.1037/0003-066X.61.1.27

40. Greenwald AG, Nosek BA, Banaji MR. Understanding and using the Implicit Association Test: I. An improved scoring algorithm. J Pers Soc Psychol. 2003;85(2):197–216. doi:10.1037/0022-3514.85.2.197

